# Beyond Inflammation: Clinical Meaning of Slow Erythrocyte Sedimentation

**DOI:** 10.64898/2026.08.26.26360269

**Authors:** Alexis Darras, Min Qiao, Kevin Peikert, Anne Hecksteden, Thomas John, Hannes Glaß, Emeric Stauffer, Ingrid Muniansi, Benoit Champigneulle, Aurelien Pichon, Michael Furian, Ivan Hancco Zirena, Julien V. Brugniaux, Alzbeta Mühlbäck, Michael J. Simmonds, Elie Nader, Philippe Joly, Tim Meyer, Samuel Verges, Andreas Hermann, Adrian Danek, Philippe Connes, Christian Wagner, Lars Kaestner

**Affiliations:** University of Bristol, United Kingdom; Saarland University, Germany; Translational Neurodegeneration Section “Albrecht Kossel”, Department of Neurology, University Medical Center Rostock, University of Rostock, Rostock, Germany; Center for Transdisciplinary Neurosciences Rostock (CTNR), University Medical Center Rostock, Rostock, Germany; United Neuroscience Campus Lund-Rostock (UNC), Rostock, Germany; University of Innsbruck, Austria; University of Lyon, France; Univ Grenoble Alpes, Inserm, CHU Grenoble Alpes, HP2, 38000 Grenoble, France; MOVE laboratory, UR 20296, University of Poitiers, France; University Hospital Zürich, Switzerland; Centro de Investigaciòn en Medicina de Altura (CIMA) Facultad de Medicina Universidad de San Martin de Porres, Peru; Huntington-Zentrum Süd, kbo Isar-Amper-Klinikum, Taufkirchen, Germany; Biorheology Research Laboratory, School of Pharmacy and Medical Sciences, Griffith University, Gold Coast, Australia; German Center for Neurodegenerative Diseases (DZNE), Rostock/Greiufswald, Rostock, Germany; Neurologische Klinik und Poliklinik, LMU Klinikum, LMU München, Munich, Germany; University of Luxemburg, Physics and Materials Science Research Unit, 1511 Luxembourg, Luxembourg

**Keywords:** erythrocyte sedimentation rate, acanthocytosis, gravitational gel collapse

## Abstract

The erythrocyte sedimentation rate (ESR) is one of the most widely used laboratory diagnostic parameters in the preliminary assessment of inflammation; indeed, every reader of this work has likely received an ESR assessment in their lifetime. A rapid ESR is a non-specific parameter that provides information about the inflammatory process. Although the origins of this methodology date back to antiquity, the prevailing view that ESR simply reflected particle settling of erythrocytes has recently undergone a paradigm shift: once cell aggregates form a system-spanning network, gravitational collapse of a weak and percolating gel reveals a more complex process reflecting the failure. The apparent non-specific nature of the cells and proteins involved also called into question the medical utility of ESR, at least in well-resourced environments. Here we show a new experimentally derived and physically modelled approach (“supraESR”) that enhances the value and accuracy of ESR for a variety of conditions that exhibit abnormally slow ESR (e.g., sickle cell disease, neuroacanthocytosis syndromes, chronic mountain sickness). We introduce a completely new diagnostic parameter that is based on an established and easily automated measurement method that promises low-cost screening for neuroacanthocytosis syndrome, a group of rare neurodegenerative diseases that are currently detectable only through integration of complex multimodal findings.

## 1 Introduction

The observation that erythrocytes settled at a faster rate in those who were ill was observed by the ancient Greeks, although it was only in the late 1800s that modern medicine attempted to quantify the erythrocyte sedimentation rate (ESR) in clinical populations[2–5]. The most common approach in current practice is remarkably simple, and involves filling a vertical tube (height 200 mm × inner diameter2.55 mm) with whole blood and measuring the distance that the erythrocyte-plasma interface migrates over (usually) 1 h. This process reduces the collective dynamics of blood sedimentation to a macroscopic reading, measured by eye and expressed in cm/h [6]. This parameter provides a non-specific marker for inflammation where greater migration per hour is associated with a proinflammatory state, and owing to its simplicity and cost-effectiveness remains a widely adopted test; usage rates reach 10 tests per 1000 inpatient days and emergency departments may request 30 ESR per 1000 visits [7, 8]. Thus while accelerated sedimentation is well understood to reflect inflammatory processes, the determinants of slow ESR remain poorly understood and without diagnostic thresholds [5] nor lower limits [6]. Classical particle sedimentation predicts a constant settling velocity once terminal velocity is achieved in a low Reynolds regime; by contrast RBC settling is intrinsically dynamic, being neither constant nor monotonic, with a striking hematocrit dependence [9, 10]. When hematocrit is above 25%, we demonstrated that erythrocytes form a percolating network that ages and collapses in a time dependent and non-linear manner [11–14]. Indeed, our advanced imaging approach employing blood with physiologically-relevant hematocrit, indicated that this percolating network is also responsible for the sharp RBC-plasma interface requisite for classic ESR measurements. When hematocrit of blood is less than 25%, however, the ability for RBC to form networks falls below a percolation network, producing diffuse interface, owing to a continuous gradient of erythrocyte concentration [1, 15, 16] (Fig. 1(a,b)). Interestingly, for these low-hematocrit samples which are common in certain anemias, regional networks of RBC may form with sufficient time and settling once local hematocrit exceeds this ~25% threshold, allowing a front of sedimentation to once again be revealed which may subsequently exhibit the network collapse mechanism. These contemporary observations support that the macroscopic and time dependent sedimentation curve reflects the microscopic properties of the percolating network, providing substantially more mechanistic understanding than classic interpretations of sedimentation, and thus also promote an opportunity for extended interpretation of ESR [12–14] (Fig.1(c,d)). In such cases, it is clear that the dynamics and underlying physical process of gel formation is distinct, although one may still evaluate meaningful parameterization using image analysis. We demonstrated, for example, that lower ESR could be explained mechanistically in neuroacanthocytosis syndrome (NAS), a group of rare hereditary neurodegenerative diseases, not simply due to altered cell shape but due to altered collapse dynamics of the percolating RBC network [11, 17]. In this study, we collected blood samples from donors with conditions expected to exhibit reduced ESR, including elite athletes, individuals with sickle cell disease, and high-altitude residents, together with those with neurodegenerative disorders including Huntington’s disease (HD), Parkinson’s disease (PD), and amyotrophic lateral sclerosis (ALS). Collectively, these diverse groups facilitated the evaluation of whether the time-dependent sedimentation phenotype observed in NAS is distinct, and thus could be used for differential diagnosis, or whether it reflected more broadly altered RBC settling mechanics. Our findings demonstrate that monitoring the interface position over time enables the extraction of more informative and accurate parameters that describe the fundamental physical properties of the blood and thus provide greater discriminatory power when compared with classic ESR parameters. This time-resolved approach offers an effective enhancement in diagnostic power of the ESR test while retaining its strengths of low cost, simplicity, and therefore accessibility.

**Fig. 1.**
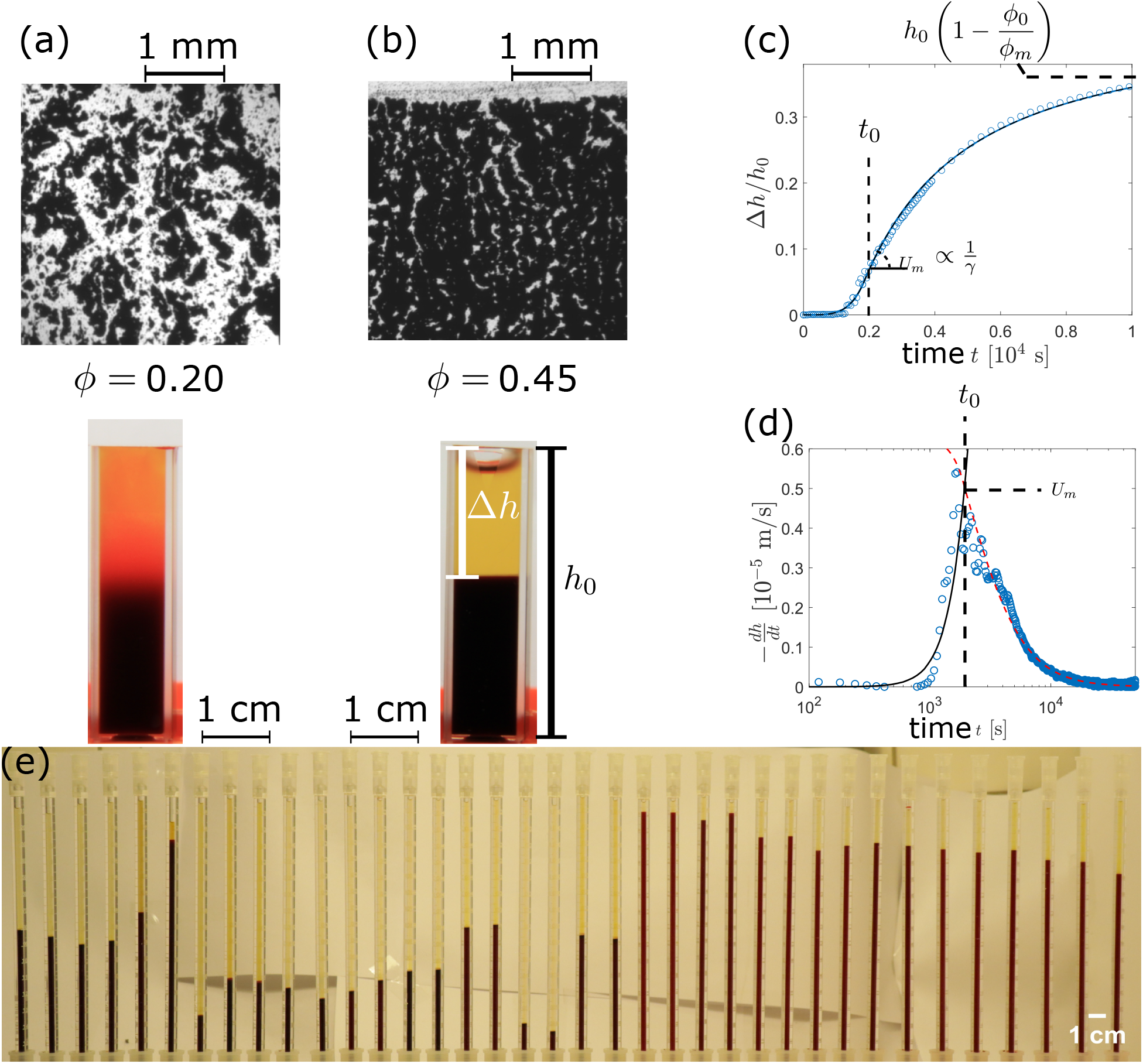
Illustration of the two possible physical regimes for erythrocyte sedimentation. (*a*) Disparate aggregates observed at hematocrits below 25 %, leading to a blurred interface, whose position determination requires some arbitrary criteria. Displayed samples have a hematocrit of 20 %, and are observed either with blue light transmission through a thin sample (top) or macroscopically (bottom) [1]. (*b*) Percolating network observed at hematocrits above 25 %, leading to a sharp interface. Displayed samples have a hematocrit of 45 %, and are observed either with blue light transmission through a thin sample (top) or macroscopically (bottom) [1]. (*c*) Annotated macroscopic sedimentation curve, observed for a 45 % hematocrit. The height of the cell-free plasma column Δ*h*, relative to the initial height of blood column *h*_0_ is plotted as a function of time *t*. The delay time *t*_0_, at which the sedimentation reaches its maximal speed *U*_*m*_, is indicated by the dashed line. This speed *U*_*m*_ depends on a dimensionless structural parameter *γ*, as well as the maximum compaction *ϕ*_*m*_. The maximal compaction *ϕ*_*m*_ also determines the final value reached by the sedimentation height Δ*h*, for a given initial height *h*_0_ and hematocrit *ϕ*_0_. Associated instantaneous sedimentation speed, fitted and annotated with relevant model parameters (see main text). (*e*) Subseries of standard Westergren tubes used to measure the ESR curves in this study.

## 2 Methods

### Study Participants and Ethical Compliance

All participants volunteered to enrol in the study via clinical and research institutions across Germany, France, and Peru. Prior to participation, study procedures and purpose were explained to volunteers in their native language, before written and informed consent was obtained. Standardized protocols were employed across all collection sites to ensure consistency in safety and data integrity. All procedures were reviewed and approved by the respective ethics committees for the specific cohorts (details provided at the end of this document).

### Study Population: Inclusion and Exclusion Criteria

Participants were adults who provided informed consent and had a confirmed clinical diagnosis corresponding to one of the target groups: Neuroacanthocytosis Syndromes (NAS), Amyotrophic Lateral Sclerosis (ALS), Parkinson’s disease (PD), Huntington’s disease (HD), Sickle Cell Disease (SCD), or who were identified as high-performance athletes or high-altitude residents. Blood samples were included only if they could be processed within the defined post-withdrawal time frame. Participants were excluded if they were experiencing an acute infection at the time of blood collection. For the analysis, the samples were divided in nine populations, defined as follows:

- **NAS**: Patients with Neuracanthocytosis syndromes (ESR independent sample size *N* = 89), defined as two core disorders being chorea-acanthocytosis (ChAc, *VPS13A* disease, OMIM #200150, *N* = 75) and McLeod syndrome (MLS, *XK* disease, OMIM #300842, *N* = 14). The diagnosis of ChAc and MLS was based on the clinical phenotype and was confirmed by detection of the *VPS13A* mutation and/or absence of chorein via Western blot (ChAc) or the detection of *XK* mutations (MLS). As no relevant significant differences were observed in their ESR (see Supplementary Material [18]), we grouped both subpopulations for subsequent analyses as NAS. Given the rarity of this disease, samples from the same patient were regarded as independent if at least one of the following condition applied: (i) samples were collected at least 1 year apart; (ii) the patient showed a clear progression in the disease; or (iii) the patient had a distinctly different medication.
- XK_prodromal_: One carrier of an *XK* mutation in the prodromal stage was collected and characterized *a posteriori*. Although not included in the statistical analyses, this sample demonstrates the ability of the present protocol to detect prodromal patients.
- **SCD**: Those with sickle-cell disease, an inherited blood disorder caused by a mutation in the hemoglobin gene, leading to the production of abnormal, sickle-shaped red blood cells that can block blood flow, cause pain, and result in progressive organ damage [19, 20] (ESR independent sample size *N* = 46).
- **Highlanders** and **CMS**: Inhabitants of La Rinconada, Peru, who lived at 5100 − 5300 m of altitude were defined as highlanders. Samples (ESR independent sample size *N* = 59) were collected during “Expedition 5300” [21]. Highlanders were differentiated as *healthy* highlanders (Highlanders; ESR independent sample size *N* = 26) or with *chronic mountain sickness* (CMS), using 2005 Consensus [22] (CMS; ESR independent sample size *N* = 33). It is worth noting that, according to a previous study, the physical properties of their blood samples mainly differ in their hematocrit, with the CMS population exhibiting a higher hematocrit (*ϕ*_0_ = 0.70 ± 0.06) than the healthy highlanders (*ϕ*_0_ = 0.58 ± 0.04)[23].
- **Sport**: Volunteers were recruited from among elite national representatives attending the Sport Medicine Department of Saarland University (ESR independent sample size *N* = 14). Blood was drawn during a regular checkup by their respective medical officers. The ESR of elite athletes is known to be lower than from healthy controls, and correlates with their fibrinogen levels, which are usually lowered [24, 25].
- **ALS**: Volunteers with Amyotrophic Lateral Sclerosis (ESR independent sample size *N* = 8).
- **HD**: Patients with Huntington’s Disease (ESR independent sample size *N* = 64).
- **PD**: Volunteers with Parkinson’s Disease (ESR independent sample size *N* = 12).
- **Control**: Healthy volunteers (ESR independent sample size *N* = 90) provided blood samples at various sites: some were collected simultaneously with NAS and served as “shipment controls” for the study, while others were collected at Saarland University.

### Sample collection and measurement

Blood samples were collected in a standardized approach at all sites. Specifically, blood was collected by qualified staff via venipuncture of the antecubital region while participants were supine and at rest. A butterfly system was used, and tourniquet time minimized, before the first volume of blood collected was discarded and then 4 mL of blood was collected into tubes containing EDTA (Becton-Dickinson, Le Pont-de-Claix, France). All tubes used for the analyses were filled to the appropriate volume and then gently mixed. Analysis was conducted as soon as practically possible at all sites. Specifically, in Germany, blood was always collected in the morning during routine patient visits and promptly transported to Saarland University in Homburg and Saarbrücken, where analysis typically began within 6 to 8 h. In France, blood was collected from those with sickle cell disease in Lyon, and then transported to Saarland University for analysis within 24 h. In Peru, blood was collected from highlanders in La Rinconada at 5100 m of altitude as part of “Expedition 5300” [21].

Measurement of ESR was conducted by transferring blood into standard Westergren tubes of 200 mm length, and then sedimentation was recorded digitally via camera (EOS M50, Canon, Tokyo, Japan) through automated remote shooting at 1 picture per minute, for a duration of at least 50 h. Up to 50 samples were recorded simultaneously (see Fig.1(e)). Measurements were performed in duplicate to minimize data loss that might occur due to tube failure, although this was not possible for sickle cell disease, owing to the smaller volume of blood available due to safety governance.

### Characterization protocol

Our approach extends the standard ESR model through continuous monitoring of the plasma-RBC interface over time. Detailed time-curves of the resultant sedimentation measurement, i.e., Δ*h*(*t*) were fitted according to our recent model [14], that is summarized as:

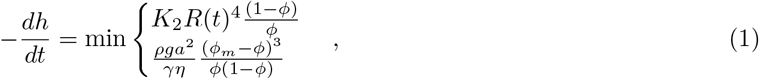

with *R*(*t*) obeying the differential equation

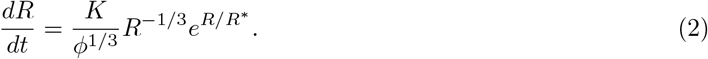

In this model, *ϕ* = *ϕ*_0_*h*_0_*/h*(*t*) is the instantaneous average volume fraction of erythrocytes within the sedimenting percolating network, i.e., the gel phase, with *ϕ*_0_ being its initial value, i.e., the sample hematocrit. The height *h*(*t*) of this gel is the complementary measurement of the usually reported sedimentation height Δ*h* = *h*_0_ − *h*, given the initial height (*h*_0_ = 20 cm for a Westergren tube). The fit parameters are *γ, ϕ*_*m*_, *K* and *K*_2_, with their initial value assessed as described previously [14]. The other parameters are fixed to their characteristic values *a* = 4 µm, *η* = 1.2 mPas, Δ*ρ* ≈ 80 kg*/*m^3^, *g* = 9.81 m*/*s^2^ and *R*^∗^ = 300 µm which we observed to be mainly constant across healthy donors [12–14]. One important secondary parameter that can be derived from the fit is the maximal (instantaneous) sedimentation speed *U*_*m*_ reached by the interface between the cell-free plasma and the RBC gel at time *t*_0_, where both lines of Eq.(1) are equal.

This parameter is quite useful, as we showed that its hematocrit dependence *ϕ*_0_ can be reliably predicted [12], enabling a normalized sedimentation speed, to be calculated for a reference hematocrit of 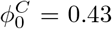 using

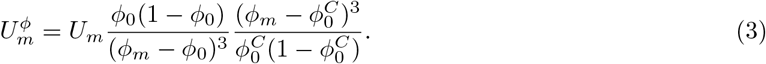

It is important to note that the previous correction assumes no change in the maximal volume fraction *ϕ*_*m*_ of the gel, and that the standard hematocrit of 0.43 was based on an average of healthy donors. Samples with deformed cells may occasionally exhibit lower final compaction *ϕ*_*m*_, which should be accounted for when renormalizing network porosity. Such a renormalization may be obtained by considering a reference maximal volume fraction 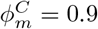 (obtained as the average of the present healthy control group):

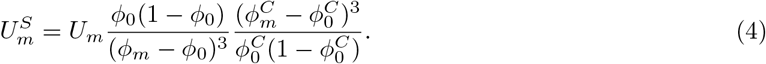

It is also possible to define a normalized maximal sedimentation speed based on the change in the maximal volume fraction *ϕ*_*m*_ of the gel, without correction for hematocrit *ϕ*_0_. This parameter is useful as it isolates the physical variable underlying distributions of sedimentation-speed *U*_*m*_. The renormalized sedimentation speed is therefore given by

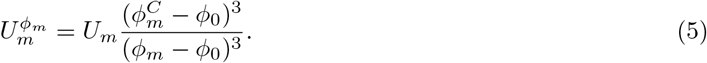

We will show in our Results section that renormalization is useful to understand the determinants of lower sedimentation rate which profoundly extends the current ESR interpretation, and the ability to identify NAS using this approach.

Kinetic characterization of the process allows an accurate and detailed description of erythrocyte sedimentation. However, given standard ESR is widely adopted, we sought to derive an equivalent parameter with similar units to enhance clinical utility. Herein, we present the “supra-ESR” value Δ*h*^*S*^ which is defined as Δ*h*^*S*^ = *U*_*m*_Δ*t*. This parameter implements our robustly measured maximum sedimentation speed *U*_*m*_ and a simple observation time Δ*t* = 1*h*, thus providing a value analogous to standard ESR output. We thus present a parameter based on the theoretical distance erythrocyte sediment once they achieve their maximal sedimentation speed, which compared with the standard ESR, provides enhanced accuracy and higher predictive power.

### 2.1 Statistical Analysis

For all extracted parameters, differences among group distributions were first assessed using Kruskal-Wallis test; *p <* 0.0001 was calculated for all parameters. Dunnett’s multiple comparison test was thus subsequently performed for all groups. For all Figure panels, p-values are symbolized as follows: n.s., not significant (*p*-value*>* 0.05); ^∗^ *p* ≤ 0.05; ^∗∗^ *p <* 0.01 and ^∗∗∗∗^ *p <* 0.0001. Effect size `e` was calculated relative to the control group using median differences, normalized by the median value of the control group To evaluate the diagnostic performance of the proposed classification criterion based on the joint distribution of 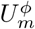 and *ϕ*_*m*_, confusion matrices were constructed by comparing predicted classifications against the known clinical diagnoses of the study participants. False-positive and false-negative rates were then derived directly from these matrices to quantify the ability of the criterion to distinguish NAS patients from controls and other clinical populations.

## 3 Results

The traditional ESR, kinetic and parameterized sedimentation metrics, and classifier performance is presented for all groups in Fig. 2. The traditional ESR did not exhibit a significantly lower ESR for SCD. Among all other groups, the athletic population (sport) exhibited the least differentiated ESR compared with controls, as emphasized by the smaller absolute effect size (`e` = 0.79) than all other groups `e` ≥ 0.88. (Fig.2(a)). In contrast, the newly-described “supra-ESR” (Fig.2(b)), produced greater differentiation among groups. This parameter derived using the maximal instantaneous speed *U*_*m*_ (Fig.2(d)), provided improved statistical differences, particularly for effect size, except for the Highlanders. In summary, these parameters demonstrated that RBC from all populations sedimented more slowly than controls.

**Fig. 2.**
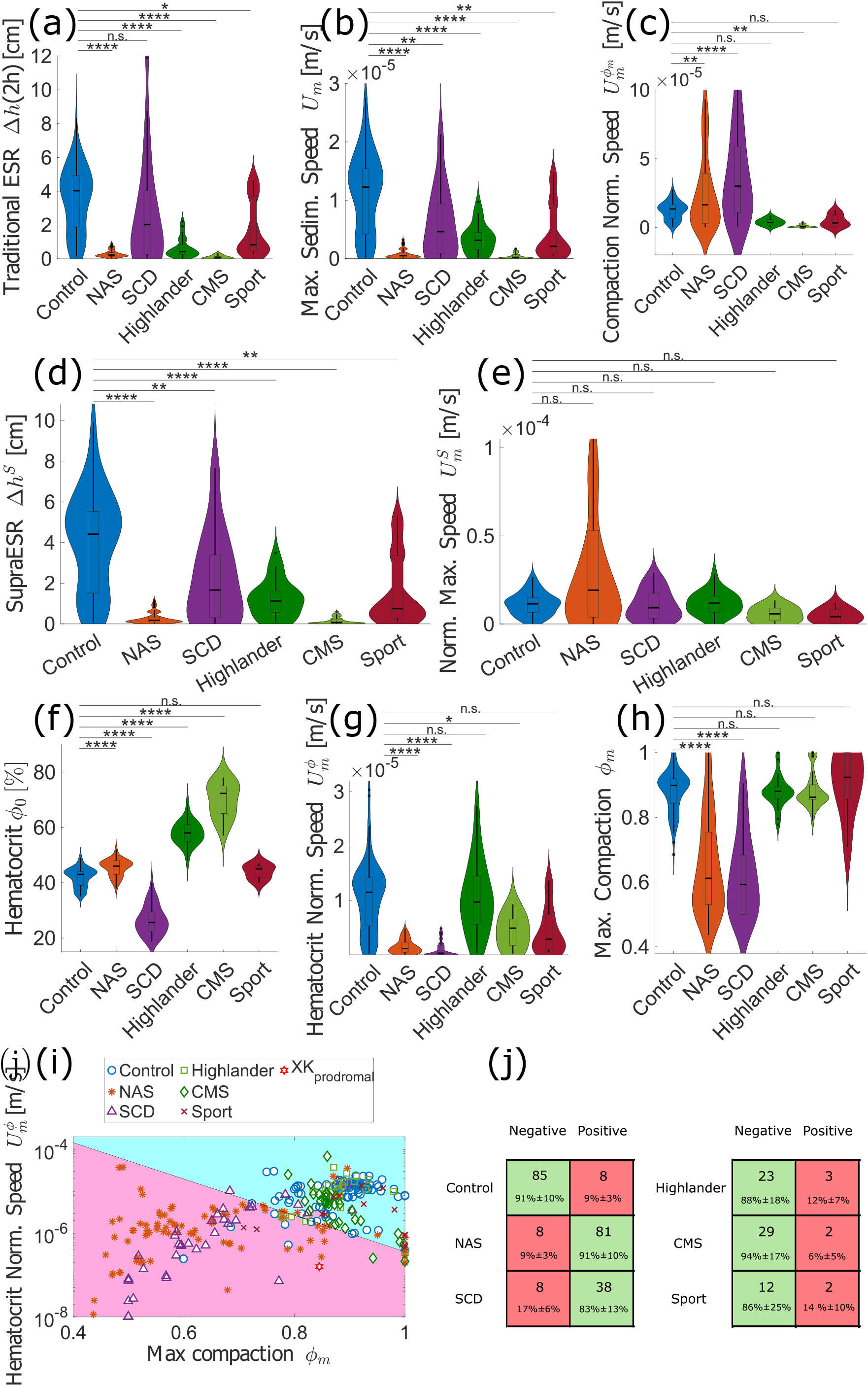
Overview of ESR metrics for populations with reduced sedimentation. (*a*) Traditional ESR after 2 h; only SCD samples show no significant decrease. (*b*) Distribution of maximal sedimentation speed *U*_*m*_. (*c*) Normalized *U*_*m*_ using Eq. (5). (*d*) Proposed “supra-ESR” Δ*h*^*S*^ = *U*_*m*_ *×* 1 h, highlighting slower ESR in all tested groups. (*e*) Normalized *U*_*m*_ via Eq. (4), removing significant differences. (*f*) Native hematocrit *ϕ*_0_; only athletes match controls. (*g*) Scaled speed 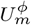 (Eq. 3); correction reduces effect size for Highlanders (from e = 0.73 to e = 0.19), CMS remains significant. (*h*) Maximal compaction *ϕ*_*m*_; SCD and NAS show markedly lower values. (*i*) 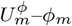 space; combined criteria separate NAS from controls (boundary: *ϕ*_*m*_ = 0.99 − 0.1 ln 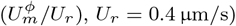. An prodromal *XK* patient (XK_prodromal_) has been added a posteriori and is also detected as positive. (*j*) Confusion matrix for this criteria.

The improved differentiation, or discriminatory power, when evaluating the instantaneous speed highlights the intrinsic features of gel collapse processes. Mechanistically, the maximal rate of sedimentation is explained by quasi-static compression,while the traditional ESR is a composite parameter describing initial gel instability and its subsequent compression. These distinct processes influence ESR according to specific and different features of blood parameters [12, 14, 15]. An advantage of instantaneous speed measurement is that it facilitates corrections using unique but physically-derived factors, including 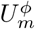 and 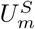 to understand the origin of deviations in Δ*h*^*S*^. In the slower sedimentation populations, for example, correcting for initial hematocrit *ϕ*_0_ and final compaction *ϕ*_*m*_, yielding the parameter 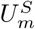, appears to explain the primary determinants of population differences in ESR (Fig.2(e)). A confirmation of this approach is evident in the Highlanders, where the effect size was reduced to zero; it is well known that blood from this group differs from healthy controls only due to increased hematocrit [23]. Collectively, these findings indicate that reduced traditional ESR from diverse groups can be explained largely through hematocrit differences, and network compaction, while the supra-ESR provides detailed gel compression dynamics.

To evaluate the specific blood properties that contribute to slower ESR, we compared the initial hematocrit *ϕ*_0_ distributions (Fig. 2(f)) with those of the control group. All populations, except for the elite athletes, differed significantly when compared with controls, although the magnitude of difference (i.e., effect size) varied. The SCD group were particularly informative, given they presented with significantly lower hematocrit, with an appreciable effect size (`e` = 0.34). This would typically infer an increased ESR, and thus the present observation that SCD is associated with slow ESR and lower supra-ESR than controls, supports that their erythrocytes have significantly altered mechanical properties that impacts gel formation and collapse dynamics. NAS patients had a mild but significant increase in hematocrit compared with controls (effect size of `e=0.07`, *<* 3% difference in median hematocrit), while healthy highlanders and CMS highlanders presented with significantly elevated hematocrit. These group differences emphasize the value in normalizing maximal sedimentation *U*_*m*_ for hematocrit (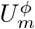, Fig. 2(g)): healthy highlanders were subsequently not different to control (`e` = 0.73 decreased to `e` = 0.19), indicating that hematocrit explained nearly all of their lower ESR. In contrast, CMS highlanders retained a lower 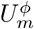 despite correction, although there was some reduction of effect size (from `e` = 0.98 to `e` = 0.59), suggesting that multiple determinants in addition to hematocrit explain low ESR in this population. It is plausible that this observation reflects that CMS highlanders present with extremely high hematocrit, and well above *ϕ*_0_ = 0.6 used in our original work as highest value to validate the hematocrit correction [12]. Indeed, given maximal compaction of erythrocytes is around *ϕ*_*m*_ = 0.9, which is very close to some of our CMS samples (*ϕ*_0_ = 0.8), it is plausible that those samples do not exhibit the characteristic initial phase of sedimentation as their erythrocyte networks may be too crowded to be fluidized prior to compression. It is acknowledged that this explanation appears to be a secondary determinant, given that, after correction for individual maximal compaction and initial hematocrit (i.e., considering 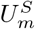), significant group differences are no longer detected. Further, we examined the maximal packing fraction *ϕ*_*m*_ (Fig. 2(h)), and found that while most populations were not different when compared with control, those with NAS and SCD presented with significantly lower packing fraction with appreciable effect sizes (|e| *>* 0.3). These later findings indicate that cell morphology and/or mechanical properties – both of which are affected in NAS and SCD – are primary contributors to maximal compaction.

Collectively, it was clear that no single parameter discriminated NAS from controls, owing largely to the significant overlap in the sample distribution. Discriminatory power was enhanced, however, through a composite of the hematocrit-corrected instant speed 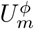 and the maximal volume fraction *ϕ*_*m*_ (Fig. 2(i)). Evaluations determined that using the criterion *ϕ*_*m*_ *<* 0.99 − 0.1 ln 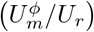, with *U*_*r*_ = 0.4 µm*/*s, NAS samples could be accurately detected from controls with less than 9 % false negative (i.e., high sensitivity) and less than 9 % false positive (i.e., high specificity). Although anecdotal, the prodromal XK_prodromal_ patient tested *a posteriori* was also classified positive using this criterion, supporting that ESR, analysed using this novel and contemporary approach, provides an accessible and cost-effective screening test for NAS disorder [11]. Of note, ~83 % of SCD samples also met this criterion. This suggests that while being highly sensitive and specific when compared with a healthy population, other disorders that reduce the deformability of erythrocytes should be ruled out in follow-up testing (e.g. smear evaluation for sickled cells). In practice, such a workflow is straightforward and remains cost-effective and accessible.

In order to test the specificity of this criterion, we also evaluated sedimentation curves from patients with non-NAS neurological disorders. Supporting the discriminatory power of ESR-based analyses, NAS was the only neurological condition that presented with a significantly decreased ESR (Fig. 3(a)). In contrast, patients with ALS or PD presented with elevated ESR, whereas ESR was not affected by HD, when compared with control. We subsequently evaluated these patients using the supra-ESR method, and found that differences between control and ALS, HD, and PD were retained (Fig. 3(b)), although the effect size was not predictably affected. These data support that ESR appears consistently elevated in several common neurological conditions while the supra-ESR is particularly sensitive to conditions, such as NAS, that are associated with abnormally low sedimentation. Factors contributing to the elevated ESR are likely plasmatic and/or hematocrit in origin. Indeed, plasma fibrinogen and other acute phase reactants, and hematocrit, are perhaps the most well-known contributors to rouleaux formation and sedimentation. These data likely explain the faster ESR for ALS in the present study given they also presented with elevated plasma fibrinogen concentration (Supp. Fig. 2(a) [18]).Further, those with PD presented with substantially abnormal hematocrit, which may explain the elevated ESR in this population (see Supp. Fig. 2(b) [18]). Intriguingly, when values were evaluated using 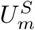 which normalized ESR values for the initial hematocrit and maximal packing fraction, the ALS and PD values remained above control (*p <* 0.05), and exhibited an appreciable effect size (e ≈ 1) (Fig. 3(c)). This supports that while hematocrit and packing fraction contribute to RBC sedimentation in these conditions, some other disease-specific processes appear meaningful and yet unknown determinants. Evaluation of the 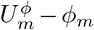 relationship (Fig. 3(d)) demonstrated that NAS was the only neurodegenerative disease to be clearly discriminated from controls by the criterion *ϕ*_*m*_ *<* 0.99−0.1 ln 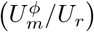, with *U*_*r*_ = 0.4 µm*/*s. However, a reasonable fraction of HD (28 %) also fulfills this criterion. This observation is biologically plausible, given erythrocytes have been demonstrated to be abnormally rigid in some HD patients [26, 27]. Nevertheless, it remains that although abnormal morphology and mechanical properties may be detected in a non-specific manner using this approach, a disease-specific signature remains elusive. In the context of HD and NAS, this is particularly important given that both present with clinically similar symptoms, making their differentiation diagnosis essential for enhanced management.

**Fig. 3.**
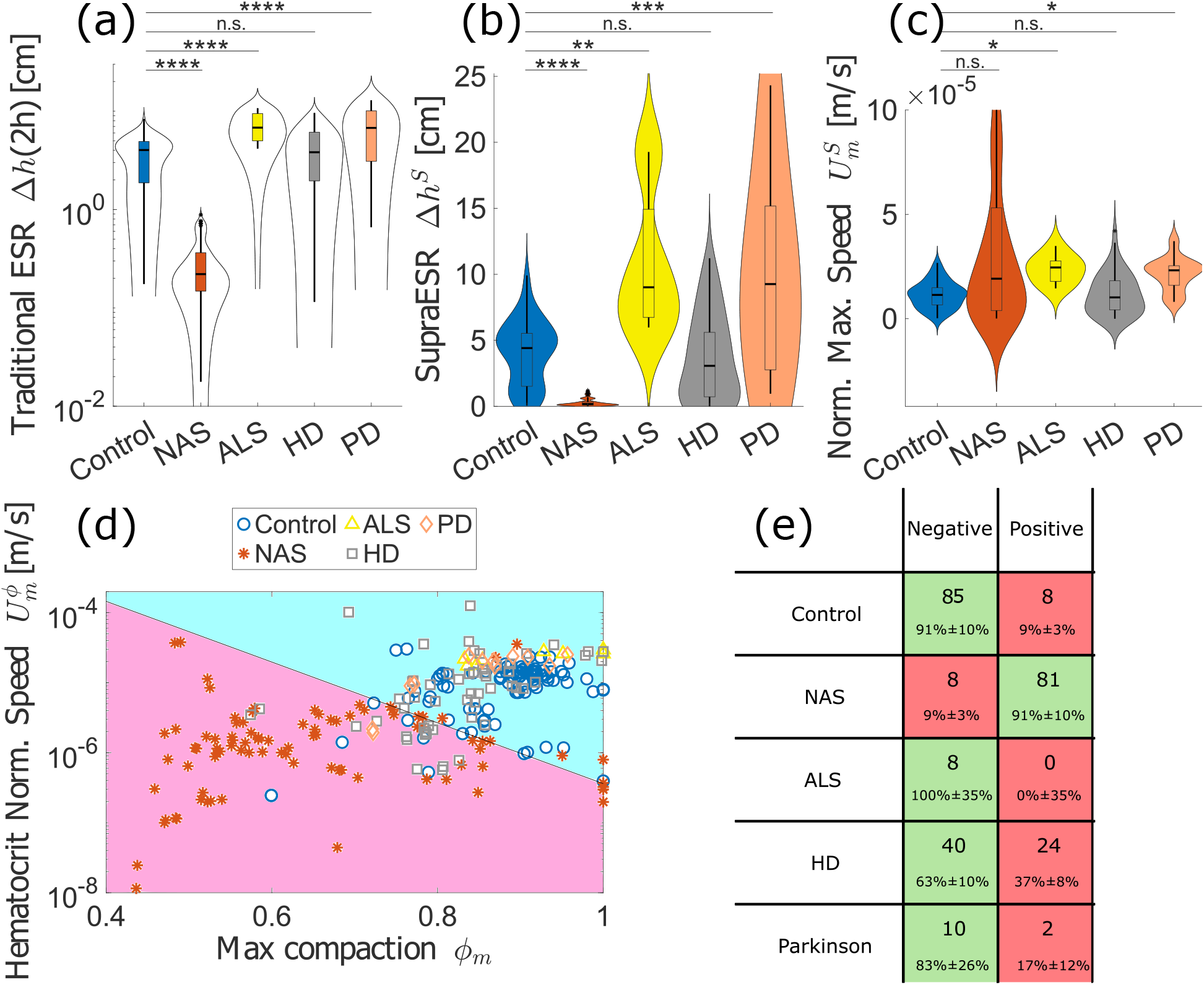
Comparisons of patients with neurological diseases. (*a*) Classical ESR value measured after 2 h. Only the NAS patients have a significantly slower ESR, ALS and PD are associated with a higher classical ESR. (*b*) Proposed “supra-ESR” Δ*h*^*S*^, which reduces the significance of all differences to the control group, except for the NAS distribution. Interestingly, the effect size is not significantly reduced, and even increased for other populations as well, highlighting that the ESR might still be influenced in other neurological diseases and that the “supra-ESR” is more sensitive to these elevated ESR. (*c*) Normalized maximum sedimentation speed 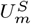 (Eq.(4)), for which all significance levels are *p >* 0.01, with the ALS and PD distribution being above the control group (*p <* 0.05, and an appreciable effect size e ≈ 1). (*d*) The same criterion as in Fig.2(i) still distinguishes the NAS population from the others. The magenta area highlights samples meeting the criteria isolating the NAS from the control population. The cyan highlights the ones grouped with the control samples.(*e*) Confusion matrix for this criteria.

## 4 Discussion

Classic ESR measurement remains an entrenched clinical biomarker owing to its accessibility more than a century after it was first introduced to clinical diagnostics. Nevertheless, its non-specific nature is limiting largely to its dependence on two successive but ultimately discrete physical processes. The first phase, reflecting the initial instability of erythrocyte gels, leads to a characteristically delayed maximal sedimentation rate, only after which the second phase of quasi-static compaction may be observed. That different physical properties of erythrocyte and plasma contribute to these independent processes leads to challenges in interpretation when using the traditional single endpoint parameter. The salient findings of this study thus indicate that the supra-ESR, introduced in this study, offers a model-based alternative that provides discrete evaluation of the independent determinants of ESR, and also enables unique normalization for the inevitable differences in underlying physical properties of blood. This approach overcomes the inherent variability that occurs between the initial gel formation and subsequent quasi-static compression (e.g., when “streamers” are formed [12, 14]), through the provision of a robust and predictive metric. Specifically, Δ*h*^*S*^ was shown to be a robust parameter with improved predictive power for distinguishing diverse populations that present with abnormal ESR. These findings significantly extend the value of ESR, and retain the cost-effectiveness per sample, relying on only modest additional hardware for analysis and interpretation. It is thus suggested that supra-ESR presents an attractive alternative for classic ESR measurement.

In addition to the direct benefits of measuring Δ*h*^*S*^, our approach enhanced discrimination of samples presenting with poor cellular deformability. A hallmark feature of SCD, beyond the classic morphological phenotype, is the profound reduction in cellular deformability – an attribute shared with NAS. The present study demonstrates the value of a new parameter derived from a composite of hematocrit-normalized maximum sedimentation speed 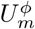 and the maximal compaction *ϕ*_*m*_. The former has clear utility for isolating the effects of hematocrit dependency (a primary determinant of blood viscosity, rouleaux formation, and ESR), which increases ESR rate in SCD. The latter, which reflects the displacement between initial and final position of the erythrocyte-plasma interface, captures the packing efficiency of the erythrocyte phase – which is sensitive to erythrocyte morphology and mechanical properties. Our newly described parameters 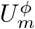 and *ϕ*_*m*_, thus logically provided discriminatory power for separating SCD and NAS from most other populations, likely due to inherent cellular morphological and mechanical impairments.. The NAS population specifically was especially separated from control values even using a simple logarithmic threshold (*ϕ*_*m*_ *<* 0.99 − 0.1 ln 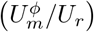), which although also identified a subset of HD and SCD samples, is consistent with known alterations in cell morphology and deformability [26–28]. The value of these parameters was further emphasized when analysis of a rare blood sample from a prodromal NAS patient XK_prodromal_ − a notoriously diagnostically challenging syndrome – was positively detected using our newly developed criteria. Although larger cohorts would strengthen this evidence, owing to its ultra-rarity, we suggest that the supra-ESR method provides a cost-effective functional screen that may identify candidates that warrant targeted diagnostic evaluation.

A particular strength of the current findings is the convergence between traditional statistical analyses and an independent classifier neuronal network (see Supplementary Material). The close alignment of discriminatory power, particularly for classifying NAS from controls, increases confidence in the present findings and is particularly valuable in this era of poor reproducibility in biomedical research [29, 30].

In summary, while the supra-ESR enhances the reliability of ESR measurements by focusing on early dynamics, the combined analysis of 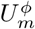 and *ϕ*_*m*_ provides a physiologically grounded and highly discriminative framework for identifying pathologically altered erythrocytes from sedimentation curves.

## 5 Conclusion

Our data show that the traditional method of collecting ESR may be extended to provide substantially more information than is captured using a single endpoint. Indeed, when the dynamics of sedimentation are captured using standard laboratory infrastructure, enhancements to the standard interface displacement (i.e., ESR 1h-point) are possible and extend to functional assessments of the mechanisms determining RBC network formation, collapse, and compaction. This extension extends a classic clinical tool from provision of non-specific marker of inflammation into a robust marker of functional readout of blood microstructure that distinguishes challenging disorders from other related pathologies. Central to our advancement is the identification of instantaneous sedimentation speed and the maximal sedimentation speed, which provided a more discriminating parameter when compared with the traditional ESR 1h-point. This classic 1h-point combines two distinct mechanisms that consist of the initial instability of gel and the subsequent quasi-static compression of RBC, while the newly-described supra-ESR approach provides discrete parameterization of these phases and thus enhances our ability to evaluate the pathology-specific determinants for each. This distinction enhances the ability to discriminate between pathologies with common blood traits, and particularly improves determination for hematocrit, plasmatic, compaction, or cell mechanical properties contributing to atypical ESR. An especially important contribution relates to our ability to evaluate changes caused by impaired cell morphology and/or mechanical properties. Our composite parameters enable the identification of the primary signature of impaired RBC packing and gel collapse, which was especially useful in identifying NAS from controls.

The implications of this work are considerable: impaired cell morphology currently requires (labor and resource intensive) microscopy studies that are subjective [26], while supra-ESR offers a cost-effective, scalable, and independent alternative that leverages an assay already embedded within many medical laboratories. The intrinsic development of measuring RBC-plasma interface dynamics offers a relatively modest financial and analytical burden, while substantially increasing the mechanistic and diagnostic value of an already highly accessible laboratory test.

Our findings enhance the physical basis of ESR interpretation – which summarizes several complex processes into a single measurement point – and rather links macroscopic blood behavior with discrete soft material properties. Our approach thus explains why traditional ESR provides generally informative, but largely non-specific, information pertaining to inflammatory states, while obscuring the discrete determinants of impaired blood behavior that is only meaningfully determined using dynamic assessments. Our work establishes supra-ESR as a quantitative extension of a classic clinical evaluation of inflammation and demonstrates predictive value even among diagnostically challenging neurological disorders.

## Supplementary information

This manuscript is accompanied by a Supplementary File, displaying the comparison of ESR values for subpopulations within NAS donors and discussing the origin of varying ESR for patients with neurological diseases, along additional analysis performed with a classifier neural network.

## Declarations

### Funding

AD acknowledges funding from the Young Investigator Grant from Saarland University. AHer is supported by the Hermann and Lilly Schilling-Stiftung. The Expedition5300 research program is supported by the Grenoble Alpes University foundation, the “Fonds de dotation AGIR pour les maladies chroniques,” and by the French National Research Agency (ANR-12-TECS-0010) in the framework of the “Investissements d’avenir” program (ANR-15-IDEX-02).

### Ethics approval and consent to participate

Blood sample collection and experiments were approved by local ethics committee, and performed after informed consent was obtained according to the Declaration of Helsinki. The main ethics approval was obtained from the “Ärztekammer des Saarlandes” (Institutional Review Board (IRB) number 51/18), while PD and ALS patients were recruited from the Technische Universität Dresden, the University Hospital of the Ludwig-Maximilian-Universität Munich and the University Medical Center Rostock (IRB # A 2019-0134). The HD patients were recruited from the University Medical Center Rostock (IRB # A 2019-0134) and from the Huntington-Zentrum Süd, kbo Isar-Amper-Klinikum (IRB # mb22061 from the “Ethik-Kommission der Bayerischen Landesärztekammer”). The NAS patients were recruited from University Medical Center Rostock (IRB # A 2019-0134), University Hospital Dresden (IRB # EK45022009 and IRB# EK517122019), and during the 11th International Meeting on Neuroacanthocytosis Syndromes in Homburg [31]. The SCD patients were treated at the University Hospital of Lyon. Samples from elite athletes were collected via the sport medicine department of Saarland university.

To analyze samples from highlanders, we conducted a cross-sectional study within the Expedition 5300 research program [21]. This study was approved by the ethics committee of the Universidad Peruana Cayetano Heredia (Lima, Peru, IRB number: 00003251) and conducted in accordance with the standards set by the Declaration of Helsinki. For SCD patients, the study was approved by the Regional Ethics Committees (L16-47, CPP Sud-Est IV, Hospices Civils de Lyon). All participants were fully informed of the study in their native language and signed a written informed consent form before inclusion.

### Data availability

All data produced in the present study are available upon reasonable request to the authors.

## Notes

### Competing Interest Statement

The authors have declared no competing interest.

### Author Declarations

Ethics committee of the Aerztekammer des Saarlandes gave ethical approval for this work (Institutional Review Board (IRB) number 51/18).

