## Supplemental Figures for "Beyond Inflammation: Clinical Meaning of Slow Erythrocyte Sedimentation"

### Too slow Erythrocyte Sedimentation Rate: Deeper biophysical understanding, novel accurate parameters and new medical applications

Alexis Darras[1], Min Qiao[2], Kevin Peikert[3-5], Anne Hecksteden[6], Thomas John[2], Hannes Glaß[3], Emeric Stauffer[8], Ingrid Muniansi[8], Benoit Champigneulle[9], Aurelien Pichon[10], Michael Furian[11], Ivan Hancco Zirena[12], Julien V. Brugniaux[9], Alzbeta Mühlbäck[13], Elie Nader[8], Philippe Joly[8], Tim Meyer[2], Samuel Verges[3], Andreas Hermann[3,4,14], Adrian Danek[15], Philippe Connes[8], Christian Wagner[2,16], Lars Kaestner[2]

[1*]University of Bristol, United Kindgom.
[2]Saarland University, Germany.
[3]Translational Neurodegeneration Section "Albrecht Kossel", Department of Neurology, University Medical Center Rostock, University of Rostock, Rostock, Germany.
[4]Center for Transdisciplinary Neurosciences Rostock (CTNR), University Medical Center Rostock, Rostock, Germany.
[5]United Neuroscience Campus Lund-Rostock (UNC), Rostock, Germany.
[6]University of Innsbruck, Austria.
[8]University of Lyon, France.
[9]Univ Grenoble Alpes, Inserm, CHU Grenoble Alpes, HP2, 38000 Grenoble, France.
[10]MOVE laboratory, UR 20296, University of Poitiers, France.
[11]University Hospital Zürich, Switzerland.
[12]Centro de Investigaciòn en Medicina de Altura (CIMA) Facultad de Medicina Universidad de San Martin de Porres, Peru.
[13]Huntington-Zentrum Süd, kbo Isar-Amper-Klinikum, Taufkirchen, Germany.
[14]German Center for Neurodegenerative Diseases (DZNE), Rostock/Greiufswald, Rostock, Germany.
[15]Neurologische Klinik und Poliklinik, LMU Klinikum, LMU München, Munich, Germany.
[16]University of Luxemburg, Physics and Materials Science Research Unit, 1511 Luxembourg, Luxembourg.

Contributing authors:;

**Abstract**

This is the supplementary material for the aforementioned publication.

**Keywords:** keyword1, Keyword2, Keyword3, Keyword4

### 1 Supplementary Figures

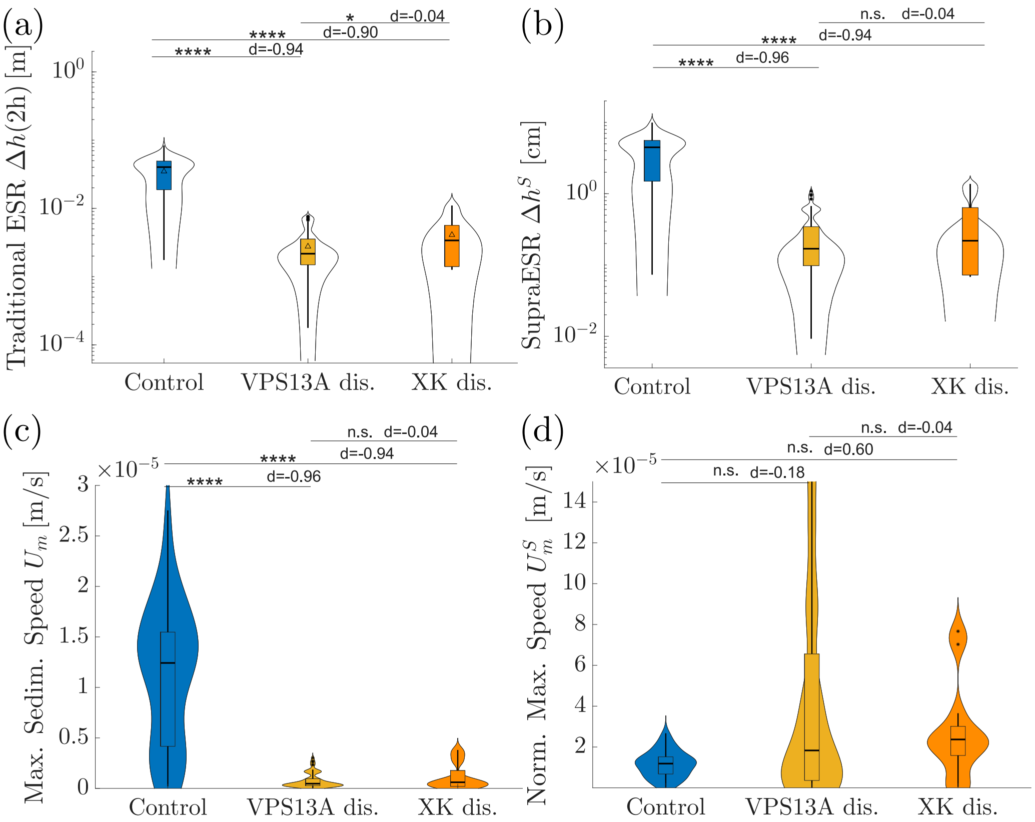


**Fig. 1** Comparison of ESR values for subpopulations within NAS donors. The diagnosis of chorea-acanthocytosis (ChAc) and McLeod Syndrom (MLS) was based on the clinical phenotype and was confirmed by detection of the with VPS13A mutation and/or absence of the erythrocyte membrane protein chorein via Western blot (ChAc) or the detection of XK mutations (MLS) (*a*) Overview of the traditional sedimentation height, measured after 2 h. (*b*) Proposed "supra-ESR" $\Delta h^{\dagger}$, which is the maximum sedimentation speed, multiplied by one hour. (*c*) The actual maximal sedimentation speed $U_m$ distribution, obtained for all populations. (*d*) Normalized maximal sedimentation speed, as obtained through Eq.(4). For all panels, significance values are obtained from a Dunnett's multiple comparison test for all populations against the control group, and symbolized as follows: n.s., not significant ($p$-value$> 0.05$); * $p < 0.05$; ** $p < 0.01$ and **** $p < 0.0001$.

### 2 Classifier Neural Network

In order to test for hidden criteria within our dataset, we also trained a Classifier Neural Network (CNN), using Matlab's dedicated Toolbox (Statistics and Machine Learning Toolbox, using the function `fitcnet`), with three fully connected layers, to classify NAS and Control samples. Data was randomly partitioned to train the network, retaining 30 % of the data as validation data, using 70 % of the data as training dataset. All fit parameters from the ESR physical model, along the haematocrit and plasma Fibrinogen level of the donors, were provided as input parameters for the CNN. The response variable was the classification as Control or NAS sample. The obtained confusion matrix is displayed in Figure 3. As the obtained accuracy is not significantly different from the one obtained by our physical criterion in the main article, one can conclude that most information is contained in the physical parameter defined through the Haematocrit normalized maximum sedimentation speed and the maximum compaction. However, as the best estimation of the accuracy seems to be slightly better with the CNN, it might be that a bigger database would lead to an even more accurate classification by a CNN using all available parameters. Attempts to train a CNN using all categories of samples (either with slow populations or neurological disorder) lead to a decreased accuracy and increased confusion, consistent with the observation that a subset of samples from SCD and HD samples are undistinguishable from the NAS population.

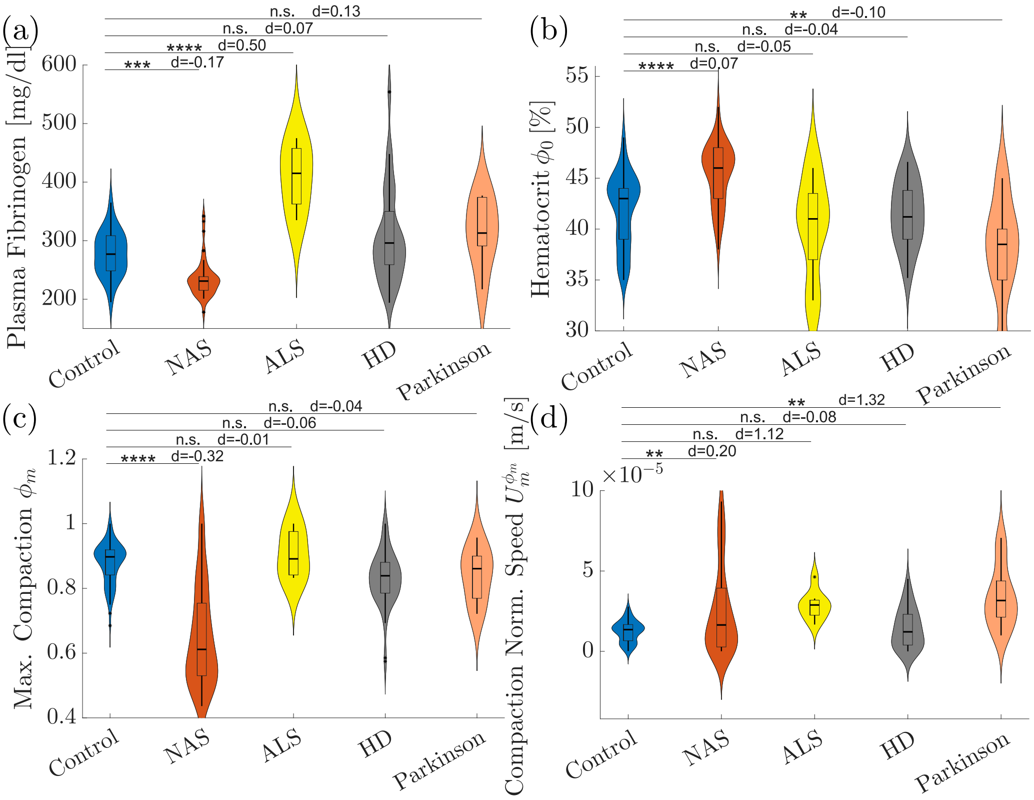


**Fig. 2** Origin of varying ESR for patients with neurological diseases. (*a*) Plasma Fibrinogen level. This parameter is significantly higher in ALS donors, with an appreciable effect size d = 0.5, which is the traditional cause for increased ESR and explains their higher ESR. (*b*) Hematocrit. Our populations of Parkinson donors has a significantly lower hematocrit, which is also associated with a higher ESR, although the effect size is moderate (d = 0.10). (*c*) Maximal compaction. Only the NAS population presents a significantly lower compaction than the control group, with an important effect size (d = −0.32). (*d*) Normalized ESR based only on the maximal volume fraction. Differences with the control group ares still significative for some populations, which indicates that this parameter is also modified by the hematocrit and fibrinogen level of the donors of these populations.

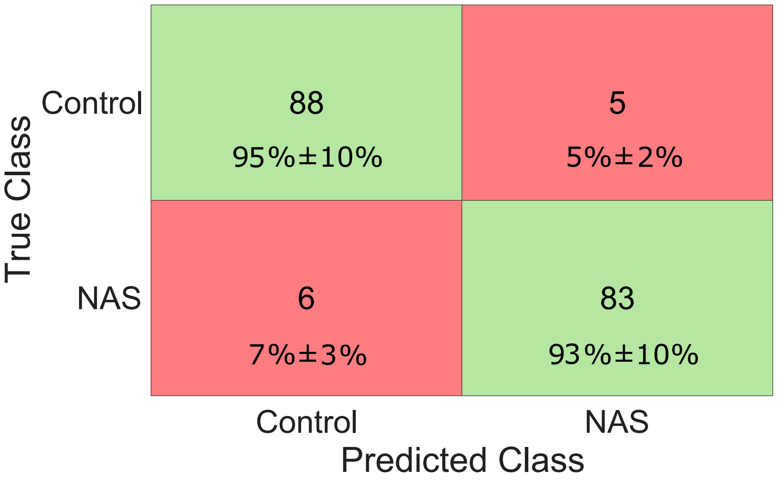


**Fig. 3** Confusion matrix of trained CNN, applied to the total dataset of Control and NAS samples.
